# Vaccine hesitancy and access to psoriasis care in the COVID-19 pandemic: findings from a global patient-reported cross-sectional survey

**DOI:** 10.1101/2022.01.20.22269546

**Authors:** K Bechman, ES Cook, N Dand, ZZN Yiu, T Tsakok, F Meynell, B Coker, A Vincent, H Bachelez, I Barbosa, MA Brown, F Capon, CR Contreras, C De La Cruz, P Di Meglio, P Gisondi, D Jullien, J Kelly, J Lambert, C Lancelot, SM Langan, KJ Mason, H McAteer, L Moorhead, L Naldi, S Norton, L Puig, P Spuls, T Torres, D Urmston, A Vesty, RB Warren, H Waweru, J Weinman, CEM Griffiths, JN Barker, CH Smith, JB Galloway, SK Mahil, On behalf of the PsoProtect study group

**Affiliations:** Centre for Rheumatic Diseases, King’s College London, London, UK; Department of Medical and Molecular Genetics, School of Basic and Medical Biosciences, Faculty of Life Sciences and Medicine, King’s College London, London, UK; Health Data Research UK, London, UK; Dermatology Centre, Salford Royal NHS Foundation Trust, The University of Manchester, Manchester Academic Health Science Centre, NIHR Manchester Biomedical Research Centre, Manchester, UK; St John’s Institute of Dermatology, Guy’s and St Thomas’ NHS Foundation Trust and King’s College London, London, UK; NIHR Biomedical Research Centre at Guy’s and St Thomas’ NHS Foundation Trust and King’s College London, London, UK; Department of Dermatology, AP-HP Hôpital Saint-Louis, Paris, France; INSERM U1163, Imagine Institute for Human Genetic Diseases, Université de Paris, Paris, France; Catedra de Dermatologia, Hospital de Clinicas, Facultad de Ciencias Medicas, Universidad Nacional de Asuncion, Paraguay; Clinica Dermacross, Santiago, Chile; St John’s Institute of Dermatology, School of Basic & Medical Biosciences, Faculty of Life Sciences & Medicine, King’s College London, London, UK; Section of Dermatology and Venereology, University of Verona, Verona, Italy; Department of Dermatology, Edouard Herriot Hospital, Hospices Civils de Lyon, University of Lyon, Lyon, France; Groupe de recherche sur le psoriasis (GrPso) de la Société française de dermatologie, Paris, France; Department of Dermatology, Ghent University, Ghent, Belgium; International Federation of Psoriasis Associations; Faculty of Epidemiology, and Population Health, London School of Hygiene and Tropical Medicine, London, UK; School of Medicine, Keele University, Keele, UK; The Psoriasis Association, Northampton, UK; Centro Studi GISED, Bergamo, Italy; Psychology Department, Institute of Psychiatry, Psychology and Neuroscience, King’s College London, UK; Department of Dermatology, Hospital de la Santa Creu i Sant Pau, Universitat Autònoma de Barcelona, Barcelona, Catalonia, Spain; Department of Dermatology, Amsterdam Public Health/Infection and Immunology, Amsterdam University Medical Centers, Location AMC, Amsterdam, The Netherlands; Department of Dermatology, Centro Hospitalar do Porto, Portugal; School of Cancer and Pharmaceutical Sciences, King’s College London, London, UK

**Author notes:** **<u>Corresponding author</u>** Dr Satveer Mahil, St John’s Institute of Dermatology, Guy’s and St Thomas’ NHS Foundation Trust, Great Maze Pond, London, SE1 9RT, UK. joint first authors (K Bechman, ES Cook). joint senior authors (JB Galloway, SK Mahil).

## Abstract

**Background:** COVID-19 vaccination is efficacious at protecting against severe COVID-19 outcomes in the general population. However, vaccine hesitancy (unwillingness for vaccination despite available vaccination services) threatens public health. Individuals taking immunosuppression for psoriasis have been prioritised for COVID-19 vaccination, however there is a paucity of information on vaccine hesitancy in this population, including contributing factors. While global healthcare has been severely disrupted in the pandemic, the impact on access to psoriasis care and whether this may negatively influence vaccine uptake, is underexplored.

**Objectives:** To explore organisational and individual factors associated with COVID-19 vaccine hesitancy in individuals with psoriasis.

**Methods:** Individuals with psoriasis, identified through global patient organisations and social media, completed a cross-sectional self-reported online survey. The primary outcome was COVID-19 vaccine hesitancy. Logistic regression was used to examine the association between predictor variables (organisational and individual factors) and outcome.

**Results:** Self-reported data from 802 individuals with psoriasis across 89 countries were available (65.6% female, median age 51 years [IQR 37-61], 43.7% taking systemic immunosuppression). Eight percent (n=63) reported vaccine hesitancy. Those reporting vaccine hesitancy were younger, more likely to be of non-white ethnicity, non-UK resident, have a lower BMI, not taking systemic immunosuppression and with shorter disease duration compared to those not reporting vaccine hesitancy. The commonest reasons for vaccine hesitancy were concerns regarding vaccine side-effects, that the vaccine is too new or that psoriasis may worsen post-vaccination. Forty percent (n=322) reported that their psoriasis care had been disrupted by the pandemic. These individuals were younger, of non-white ethnicity, with shorter duration and more severe psoriasis. Disruption to psoriasis care was associated with vaccine hesitancy (unadjusted OR 2.97 (95%CI 1.23-7.13), p=0.015), although not statistically significant in the adjusted model.

**Conclusion:** A minority of individuals with psoriasis from our study reported COVID-19 vaccine hesitancy. Similar to general population trends, vaccine hesitancy in our psoriasis sample is most common in younger age and ethnic minority groups. Our data highlight patient concerns regarding COVID-19 vaccination, which are important to address during patient-clinician interactions to help optimise vaccine uptake and mitigate risks from the ongoing pandemic in individuals with psoriasis.

**Key points:** *What’s already known about this topic?:*

- The COVID-19 vaccine is highly efficacious at protecting against severe COVID-19 outcomes in the general population. Vaccine hesitancy (unwillingness to receive vaccination despite available vaccination services) poses a major threat to global public health and is more common in women, younger age and ethnic minority groups in the general population.
- Individuals with psoriasis taking systemic immunosuppression were considered at high risk of severe COVID-19 outcomes and prioritised for vaccination, however there is a paucity of information on vaccine hesitancy in this group, including contributing factors.
- While global healthcare has been severely disrupted by the COVID-19 pandemic, access to psoriasis care and its potential impact on vaccine hesitancy is underexplored.

*What does this study add?:*

- A substantial proportion (40%) of individuals with psoriasis reported disrupted access to psoriasis care during the COVID-19 pandemic. Disrupted care was most commonly reported in younger age and ethnic minority groups.
- COVID-19 vaccine hesitancy was reported by a minority (8%) of individuals with psoriasis. Those reporting vaccine hesitancy were younger and more likely to be of non-white ethnicity, in keeping with trends in the general population.
- The commonest reasons for vaccine hesitancy were concerns regarding vaccine side effects, that the vaccine is too new or that psoriasis may worsen post-vaccination. These concerns are important to address during patient-clinician interactions to help mitigate risks from the ongoing pandemic in individuals with psoriasis.

## Introduction

The COVID-19 pandemic has had a major impact on global healthcare delivery. In the UK, the urgent need for infection control drove NHS services to rapidly redesign care delivery^1^. In primary care this involved introducing digital triage and expanding remote consultations^2,3^. In secondary care, outpatient clinics transitioned from face-to-face consultations to digital modalities, and appointments were guided by remote monitoring and patient-initiated follow-up. Global healthcare systems have also responded to the urgent need to deliver mass COVID-19 vaccine rollout programmes. The impact of these major institutional shifts on the management of patients with chronic inflammatory diseases such as psoriasis remains under-explored.

The COVID-19 vaccine has proven to be highly efficacious at protecting against severe COVID-19 outcomes in the global general population^4–6^. However, many remain unvaccinated, often due to personal choice^7,8^. Vaccine hesitancy, defined as delay in acceptance or refusal of vaccination despite availability of vaccination services, is a major threat to global public health^9^. In the UK general population, COVID-19 vaccine hesitancy is higher in women, younger age and ethnic minority groups^10–13^. Individuals with psoriasis, particularly those prescribed immunosuppressant therapies, are considered a high priority in the COVID-19 vaccination programme^14,15^. However, there is a paucity of information on vaccine hesitancy among clinically vulnerable groups including psoriasis patients, and the factors contributing to this^16^. This knowledge will help to inform immediate priorities for clinical care and mitigate risks from the ongoing pandemic.

We sought to understand perceptions around COVID-19 vaccination in individuals with psoriasis and characterise the organisational and individual factors associated with vaccine hesitancy. Specifically, we explored the impact of disrupted access to psoriasis care and demographic factors on COVID-19 vaccine hesitancy using a global patient-reported cross-sectional survey.

## Methods

### Study, Design and Participants

An online self-report survey was designed for people with psoriasis (Psoriasis Patient Registry for Outcomes, Therapy and Epidemiology of COVID-19 Infection *Me*, PsoProtect*Me*)^17^. PsoProtect*Me* was launched on 4th May 2020, disseminated globally via social media, patient organisations and clinical networks. The questionnaire was updated in May 2021 to include additional sections exploring the impact of the ongoing pandemic on access to medical care, perception of immunosuppressant-associated risks and COVID-19 vaccine uptake. Follow up questions were defined by our expert study group (clinicians, epidemiologists, health data researchers, patient representatives) and circulated to existing respondents of PsoProtect*Me*, as well as being open to new participants. Data were collected and managed using REDCap electronic data capture tools, licensed to King’s College London^18^.

### Variables

The methodology and study variables within PsoProtect*Me* are previously described^18^. For the current analysis, systemic immunosuppressant medications were classified into either standard systemic or targeted therapy. Standard systemic therapies included: acitretin, apremilast, ciclosporin, fumaric acid esters/dimethylfumarate, methotrexate (oral or subcutaneous) and prednisolone. Targeted therapies included tumour necrosis factor (TNF) (adalimumab, certolizumab pegol, etanercept, golimumab, infliximab), interleukin (IL)-17 inhibitors (brodalumab, ixekizumab, secukinumab) and IL-12/IL-23p40 or IL-23p19 inhibitors (guselkumab, risankizumab, tildrakizumab, ustekinumab).

Access to care was assessed by asking participants to rate their agreement with the following statement: “In the pandemic, my psoriasis care has been affected because I have had problems accessing the doctors and nurses who usually care for me:” Perception of medication risk was assessed by asking participants to rate agreement with the following statements: “This medication makes me more at risk of catching COVID-19” and “This medication makes me more at risk of poor recovery from COVID-19.” A 5-point ordinal scale for each of the above statements included the following responses: “strongly agree”, “moderately agree”, “no feelings”, “moderately disagree” and “strongly disagree”.

The primary outcome was COVID-19 vaccine hesitancy, defined as unwillingness to receive the vaccine^9^. This was assessed using the following questions: “Do you plan to have the vaccine?” and “Why have you not received at least 1 dose of the COVID-19 vaccine?”. Reasons for vaccine hesitancy were evaluated using the following question: “Why did you decide not to have the vaccine? (Check all that apply). The following responses were available (designed to capture known factors driving vaccination behaviours^19^): “I do not think it will protect me from COVID-19”, “I do not think I need it as I am not at risk of serious illness from COVID-19”, “I have already had COVID-19, so I do not think I need the vaccine”, “I am worried about the side effects from the vaccine”, “The vaccine is too new for me to feel confident about it”, “It is difficult for me to get the vaccine”, “My doctor has not discussed it with me yet”, “My friends/family have advised me not to have it”, “I am worried my psoriasis will get worse after the vaccine”, “I am worried about having the vaccine because of my tablet/injection medicines for my psoriasis” and “Other (please specify)”.

### Statistical Methods

Data were extracted on 9th August 2021 and analysed using Stata 16. Descriptive statistics were used to compare differences in baseline demographics. The impact of the pandemic on access to psoriasis care and vaccination hesitancy were described and graphically presented by population subgroups, age, ethnicity and psoriasis treatment. Logistic regression was used to examine vaccine hesitancy. Disrupted access to care was analysed as ordered categorical variables with ‘strongly disagree’ as the reference group. A minimally adjusted model included the following covariates: age, sex, and ethnicity. Missing data were addressed using multiple imputation (Appendix S1). Results between complete case and imputed models were compared.

## Results

### Demographic and clinical characteristics of study participants

Self-reported data were available from 802 individuals with a primary diagnosis of psoriasis who completed questions exploring the impact of the COVID-19 pandemic on their medical care. Baseline demographic data were available for 655 (81.7%) participants (Table 1). Responses were collected from 89 countries (69% from the UK).

**Table 1.**
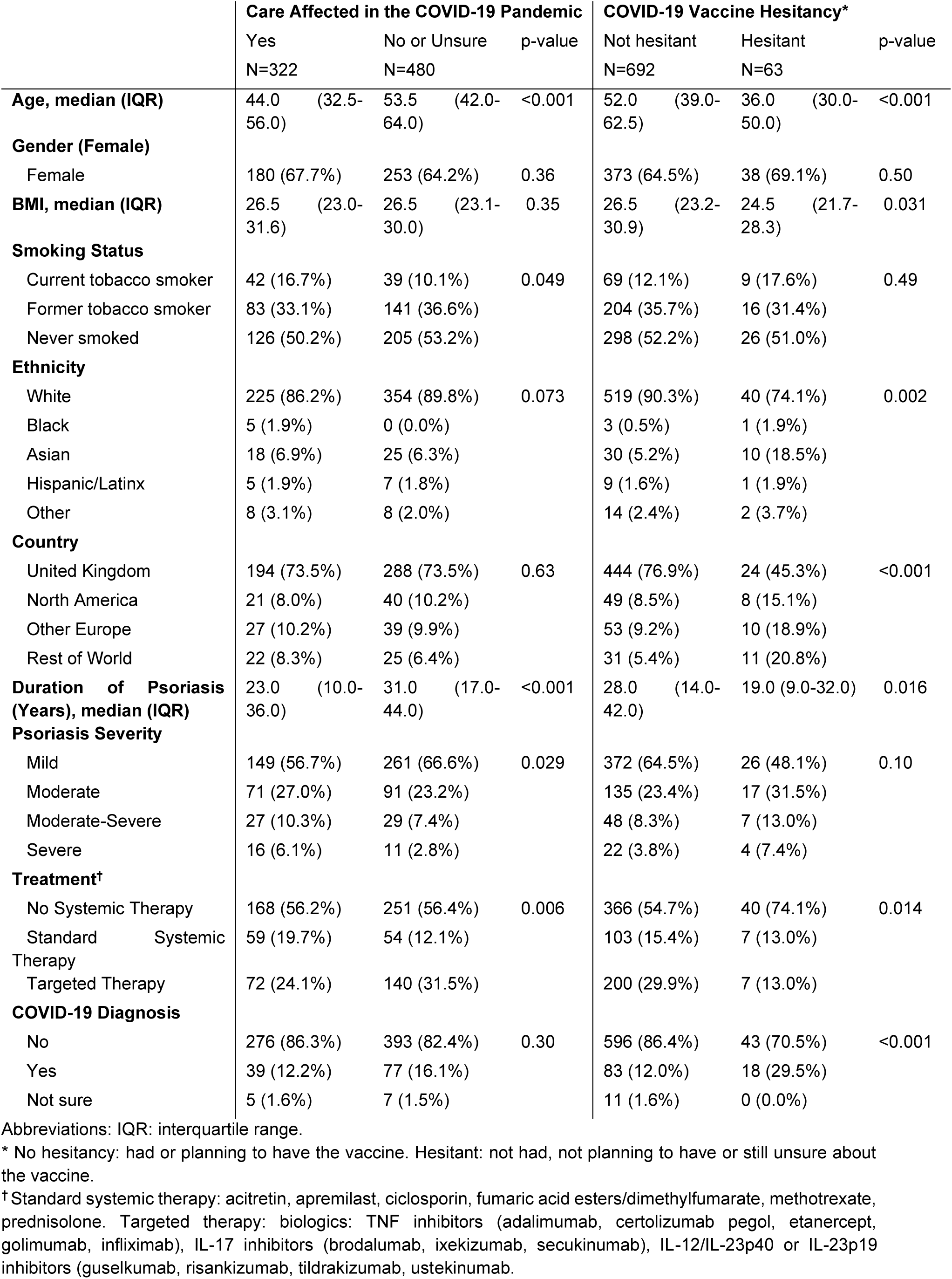
Demographics of study population.

|  | Care Affected in the COVID-19 Pandemic |  |  |  | COVID-19 Vaccine Hesitancy* |  |  |
| --- | --- | --- | --- | --- | --- | --- | --- |
|  | Yes<br>N=322 |  | No or Unsure<br>N=480 | p-value | Not hesitant<br>N=692 | Hesitant<br>N=63 | p-value |
| **Age, median (IQR)** | 44.0 (32.5-56.0) |  | 53.5 (42.0-64.0) | <0.001 | 52.0 (39.0-62.5) | 36.0 (30.0-50.0) | <0.001 |
| **Gender (Female)** |  |  |  |  |  |  |  |
| Female | 180 (67.7%) |  | 253 (64.2%) | 0.36 | 373 (64.5%) | 38 (69.1%) | 0.50 |
| **BMI, median (IQR)** | 26.5 (23.0-31.6) |  | 26.5 (23.1-30.0) | 0.35 | 26.5 (23.2-30.9) | 24.5 (21.7-28.3) | 0.031 |
| **Smoking Status** |  |  |  |  |  |  |  |
| Current tobacco smoker | 42 (16.7%) |  | 39 (10.1%) | 0.049 | 69 (12.1%) | 9 (17.6%) | 0.49 |
| Former tobacco smoker | 83 (33.1%) |  | 141 (36.6%) |  | 204 (35.7%) | 16 (31.4%) |  |
| Never smoked | 126 (50.2%) |  | 205 (53.2%) |  | 298 (52.2%) | 26 (51.0%) |  |
| **Ethnicity** |  |  |  |  |  |  |  |
| White | 225 (86.2%) |  | 354 (89.8%) | 0.073 | 519 (90.3%) | 40 (74.1%) | 0.002 |
| Black | 5 (1.9%) |  | 0 (0.0%) |  | 3 (0.5%) | 1 (1.9%) |  |
| Asian | 18 (6.9%) |  | 25 (6.3%) |  | 30 (5.2%) | 10 (18.5%) |  |
| Hispanic/Latinx | 5 (1.9%) |  | 7 (1.8%) |  | 9 (1.6%) | 1 (1.9%) |  |
| Other | 8 (3.1%) |  | 8 (2.0%) |  | 14 (2.4%) | 2 (3.7%) |  |
| **Country** |  |  |  |  |  |  |  |
| United Kingdom | 194 (73.5%) |  | 288 (73.5%) | 0.63 | 444 (76.9%) | 24 (45.3%) | <0.001 |
| North America | 21 (8.0%) |  | 40 (10.2%) |  | 49 (8.5%) | 8 (15.1%) |  |
| Other Europe | 27 (10.2%) |  | 39 (9.9%) |  | 53 (9.2%) | 10 (18.9%) |  |
| Rest of World | 22 (8.3%) |  | 25 (6.4%) |  | 31 (5.4%) | 11 (20.8%) |  |
| **Duration of Psoriasis (Years), median (IQR)** | 23.0 (10.0-36.0) |  | 31.0 (17.0-44.0) | <0.001 | 28.0 (14.0-42.0) | 19.0 (9.0-32.0) | 0.016 |
| **Psoriasis Severity** |  |  |  |  |  |  |  |
| Mild | 149 (56.7%) |  | 261 (66.6%) | 0.029 | 372 (64.5%) | 26 (48.1%) | 0.10 |
| Moderate | 71 (27.0%) |  | 91 (23.2%) |  | 135 (23.4%) | 17 (31.5%) |  |
| Moderate-Severe | 27 (10.3%) |  | 29 (7.4%) |  | 48 (8.3%) | 7 (13.0%) |  |
| Severe | 16 (6.1%) |  | 11 (2.8%) |  | 22 (3.8%) | 4 (7.4%) |  |
| **Treatment†** |  |  |  |  |  |  |  |
| No Systemic Therapy | 168 (56.2%) |  | 251 (56.4%) | 0.006 | 366 (54.7%) | 40 (74.1%) | 0.014 |
| Standard Systemic Therapy | 59 (19.7%) |  | 54 (12.1%) |  | 103 (15.4%) | 7 (13.0%) |  |
| Targeted Therapy | 72 (24.1%) |  | 140 (31.5%) |  | 200 (29.9%) | 7 (13.0%) |  |
| **COVID-19 Diagnosis** |  |  |  |  |  |  |  |
| No | 276 (86.3%) |  | 393 (82.4%) | 0.30 | 596 (86.4%) | 43 (70.5%) | <0.001 |
| Yes | 39 (12.2%) |  | 77 (16.1%) |  | 83 (12.0%) | 18 (29.5%) |  |
| Not sure | 5 (1.6%) |  | 7 (1.5%) |  | 11 (1.6%) | 0 (0.0%) |  |

Abbreviations: IQR: interquartile range.
\* No hesitancy: had or planning to have the vaccine. Hesitant: not had, not planning to have or still unsure about the vaccine.
† Standard systemic therapy: acitretin, apremilast, ciclosporin, fumaric acid esters/dimethylfumarate, methotrexate, prednisolone. Targeted therapy: biologics: TNF inhibitors (adalimumab, certolizumab pegol, etanercept, golimumab, infliximab), IL-17 inhibitors (brodalumab, ixekizumab, secukinumab), IL-12/IL-23p40 or IL-23p19 inhibitors (guselkumab, risankizumab, tildrakizumab, ustekinumab).

### Disrupted access to psoriasis care during the COVID-19 pandemic

Forty percent of participants (n=322) reported that access to their psoriasis care had been disrupted during the COVID-19 pandemic, either strongly agreeing or moderately agreeing to the question (Table 1, Figure 1). Individuals with disrupted access to care were younger (median age 44 years [interquartile range (IQR) 33-56] versus 54 years [IQR 42-64]) and more likely to be of non-white ethnicity. They had a shorter duration of psoriasis (median 23 years [IQR 10-36] versus 31 years [IQR 17-44]), likely a reflection of their younger age, and had more severe psoriasis (6.1% versus 2.8%). The proportion of participants taking systemic medications were similar between those with and without disrupted access to care. However, there were differences in the prescription of standard systemic and targeted immunosuppressant therapies, with a smaller proportion on targeted therapy in those with disrupted access to care compared to those without (55%, n=72 versus 72%, n=140).

**Figure 1.**
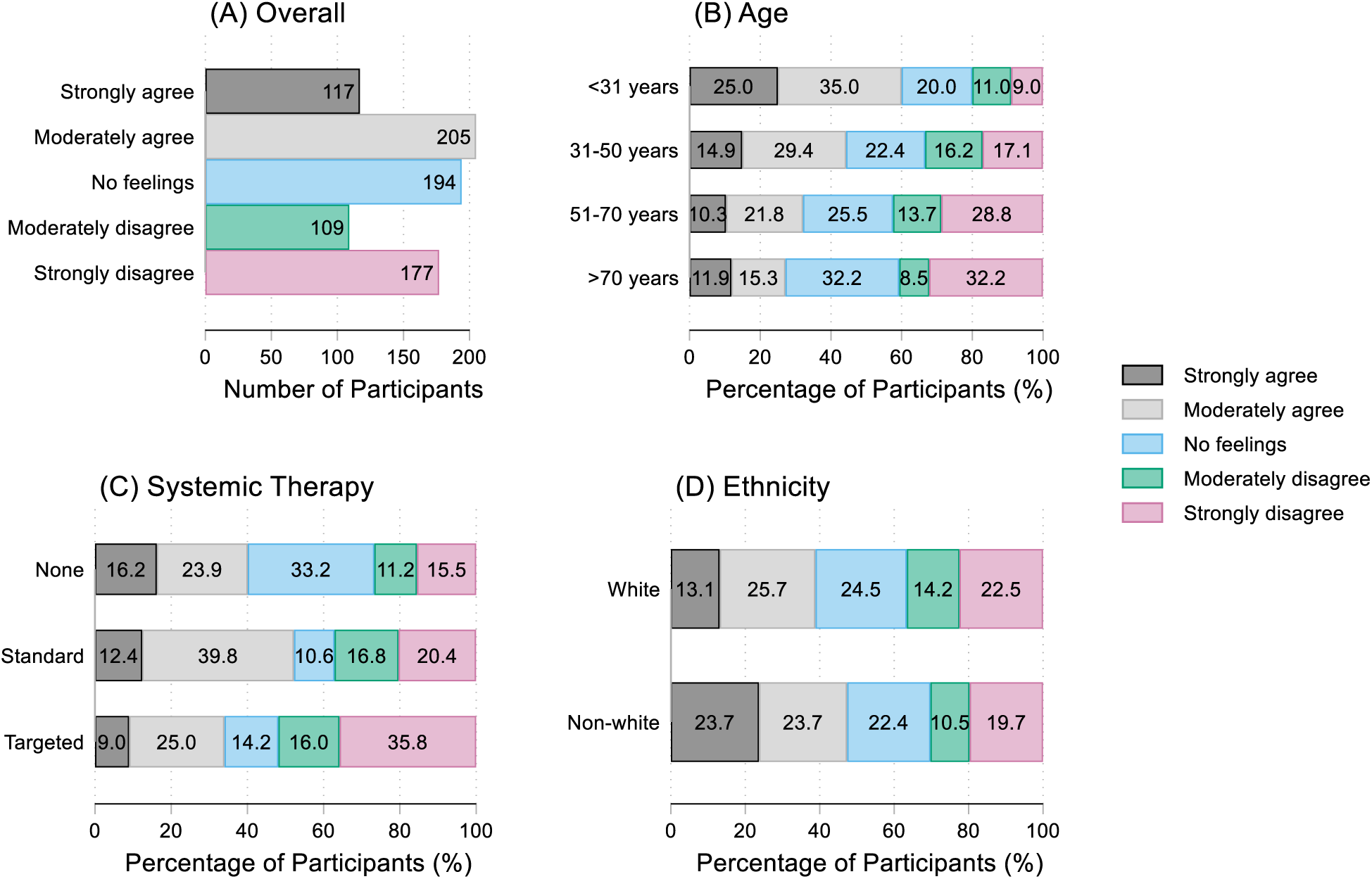
Extent to which participants feel their psoriasis care has been affected by the COVID-19 pandemic. (A) **overall count**. (B) **by age group**; <31 years: n=100, 31-50 years: n=228, 51-70 years: n=271, >70 years: n=59. (C) **by treatment type**; no systemic therapy: n=419; standard systemic therapy: n=113; targeted therapy: n=212. (D) **by ethnicity**; white: n=579; non-white: n=76.

### Systemic immunosuppressant COVID-19 risk perception and adherence

Participants were asked whether their standard systemic or targeted immunosuppressant therapy conferred an increased risk of contracting COVID-19. Of 325 individuals taking immunosuppression who answered this question, more than half (179, 55.1%) felt their medication increased infection risk, the majority of whom were on targeted therapy (116 vs. 63). More than half (183, 56.3%) also felt their medication increased their risk of poor COVID-19 recovery. There was variation in population subgroups, with a greater risk reported by those in the youngest age groups and in individuals of white ethnicity.

Participants were also asked if they adhered to their immunosuppressant medication. We excluded any individuals who stopped medication during a COVID-19 infective episode. Of those participants taking immunosuppression answering this question (n=328), 115 (35%) were prescribed standard systemic therapy and 213 (65%) were prescribed targeted therapy alone or combination targeted and standard systemic therapy. Seventeen percent of individuals in each treatment group stopped their therapy during the pandemic; (n=20 in the standard systemic group and n=37 in the targeted therapy group).

### COVID-19 vaccine hesitancy

Of 755 participants who answered questions on COVID-19 vaccine uptake, 611 (80.9%) had received at least 1 vaccine dose. Of these individuals, 99 (16.2%) reported that their psoriasis worsened following vaccination, with 63 describing changes occurring within 2 weeks.

Sixty-three participants (8.3%) had declined the COVID-19 vaccine or were not planning to have it (i.e. vaccine hesitant, Table 1 & Figure 2). Compared to those who were not vaccine hesitant, these individuals were younger [median age 36 years (IQR 30-50) vs. 52 years (IQR 39-63)], more likely to be of non-white ethnicity (20.0% vs. 7.2%), live outside the UK (13.6% vs. 5.1%), have a numerically lower BMI (median 24.5kg/m2 [IQR 21.7-28.3] versus 26.5kg/m2 [23.2-30.9]) and a shorter disease duration (median length 19 years [IQR 9-32] vs. 28 years [IQR 14-42]). They were less likely to be taking systemic immunosuppression; a smaller proportion of those reporting vaccine hesitancy were taking systemic immunosuppression (standard or targeted) compared to those who were not vaccine hesitant (26.0% versus 45.3%). The three commonest reasons for vaccine hesitancy were concerns regarding potential side effects, that the vaccine is too ‘new’ and that their psoriasis will worsen post vaccination (Figure 2).

**Figure 2.**
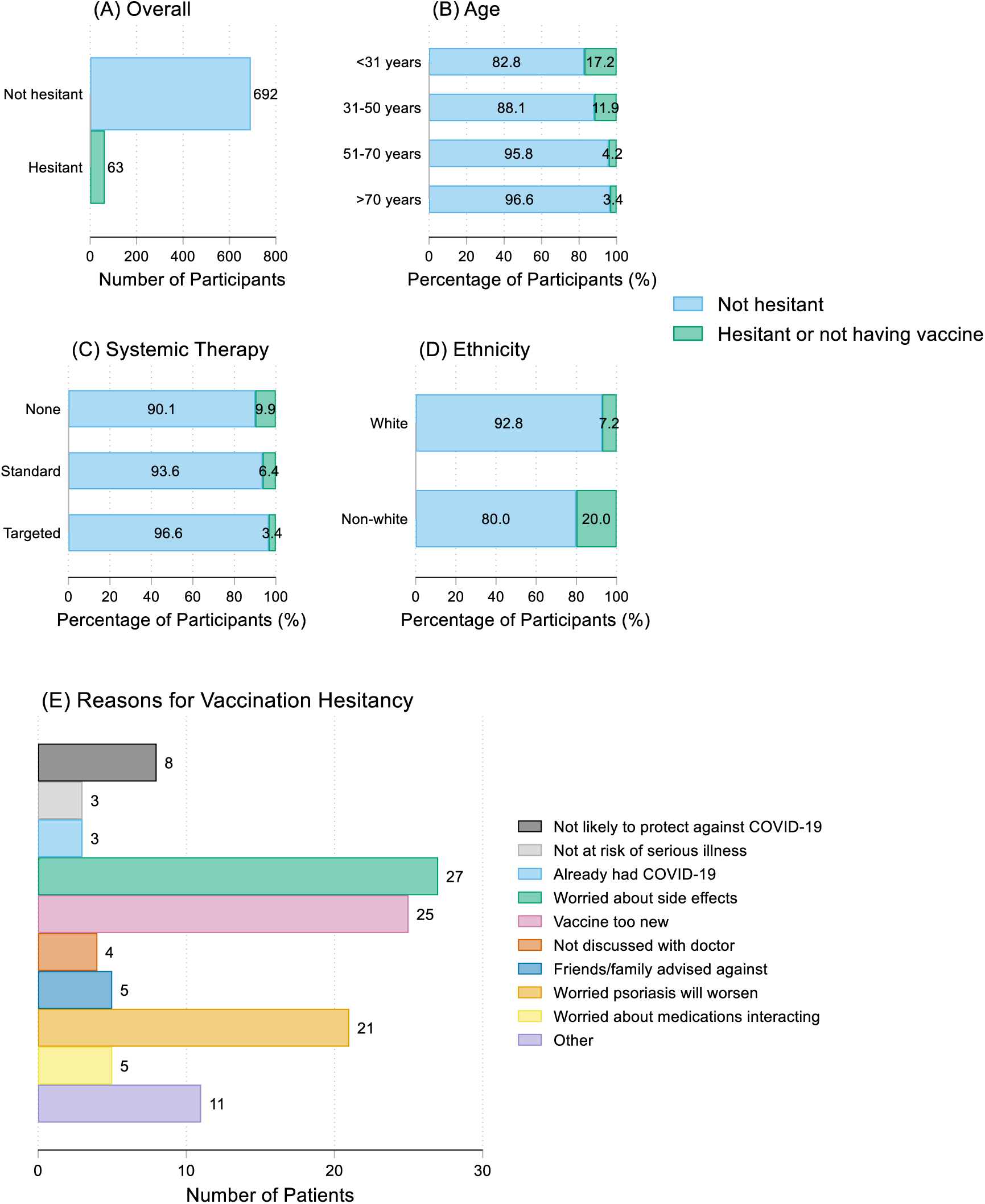
COVID-19 vaccine hesitancy. (A) overall count. (B) by age group; <31 years: n=93; 31-50 years: n=219; 51-70 years: n=261; >70 years: n=58. (C) by treatment; no systemic therapy: n=406; standard systemic therapy: n=110; targeted therapy: n=207. (D) by ethnicity; white: n=559; non-white: n=70. (E) reasons for vaccine hesitancy.

### Factors associated with COVID-19 vaccine hesitancy

A logistic regression model was used to examine the association between self-reported disrupted access to psoriasis care during the pandemic and vaccine hesitancy. In the unadjusted model, strongly agreeing that psoriasis care was disrupted was associated with vaccine hesitancy, compared to those who strongly disagreed (odds ratio [OR] 2.97, 95% CI 1.23 to 7.13, p=0.015). The direction of association remained when adjusting for age, sex, and ethnicity, although not statistically significant (adjusted OR 1.90, 95% CI 0.72 to 5.05, p=0.196) (Figure 3). Due to missing demographic data (Supplementary Table S1-S2), a multiple imputation model was fitted. In the imputed multivariate model, the association was stronger but remained non-significant (adjusted OR 2.32 [95% CI 0.94 to 5.71], p=0.068).

**Figure 3.**
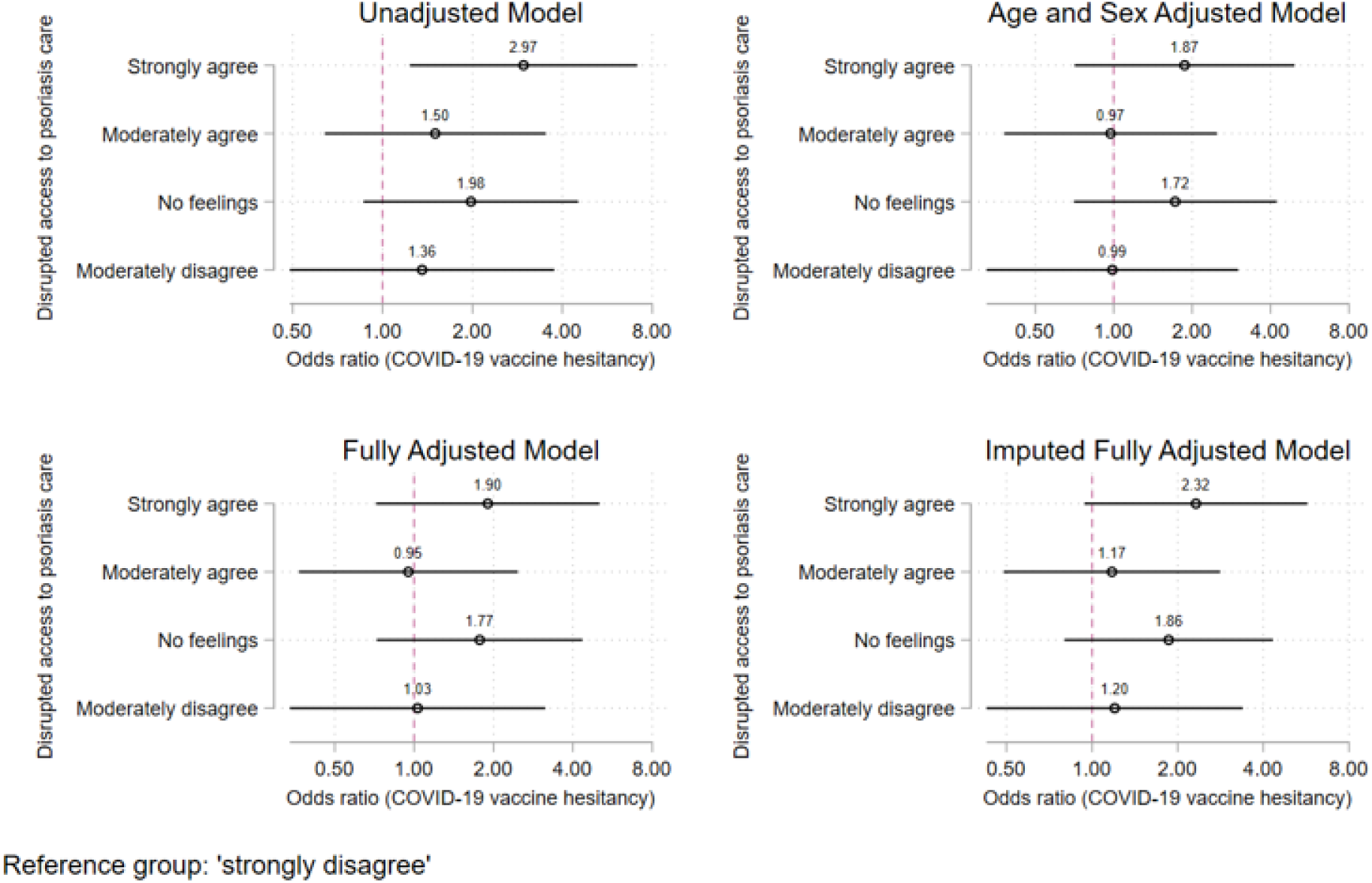
Association between disrupted access to psoriasis care and COVID-19 vaccine hesitancy.

The association between immunosuppressant medication adherence and vaccine hesitancy was also examined. This included 320 participants taking standard systemic, targeted immunosuppression or a combination of both. Of these 56 (17.5%) were not adherent to at least one of their systemic immunosuppressants. There was no evidence for an association between non-adherence and vaccine hesitancy in the unadjusted and adjusted models (unadjusted OR 1.93 [95% CI 0.58 to 6.39], p=0.28, and adjusted OR 2.96 [95% CI 0.77 to 11.3], p=0.11) (Figure 4). However, this may reflect the limited sample of individuals on systemic therapy who completed the vaccine questionnaire and warrants further investigation.

**Figure 4.**
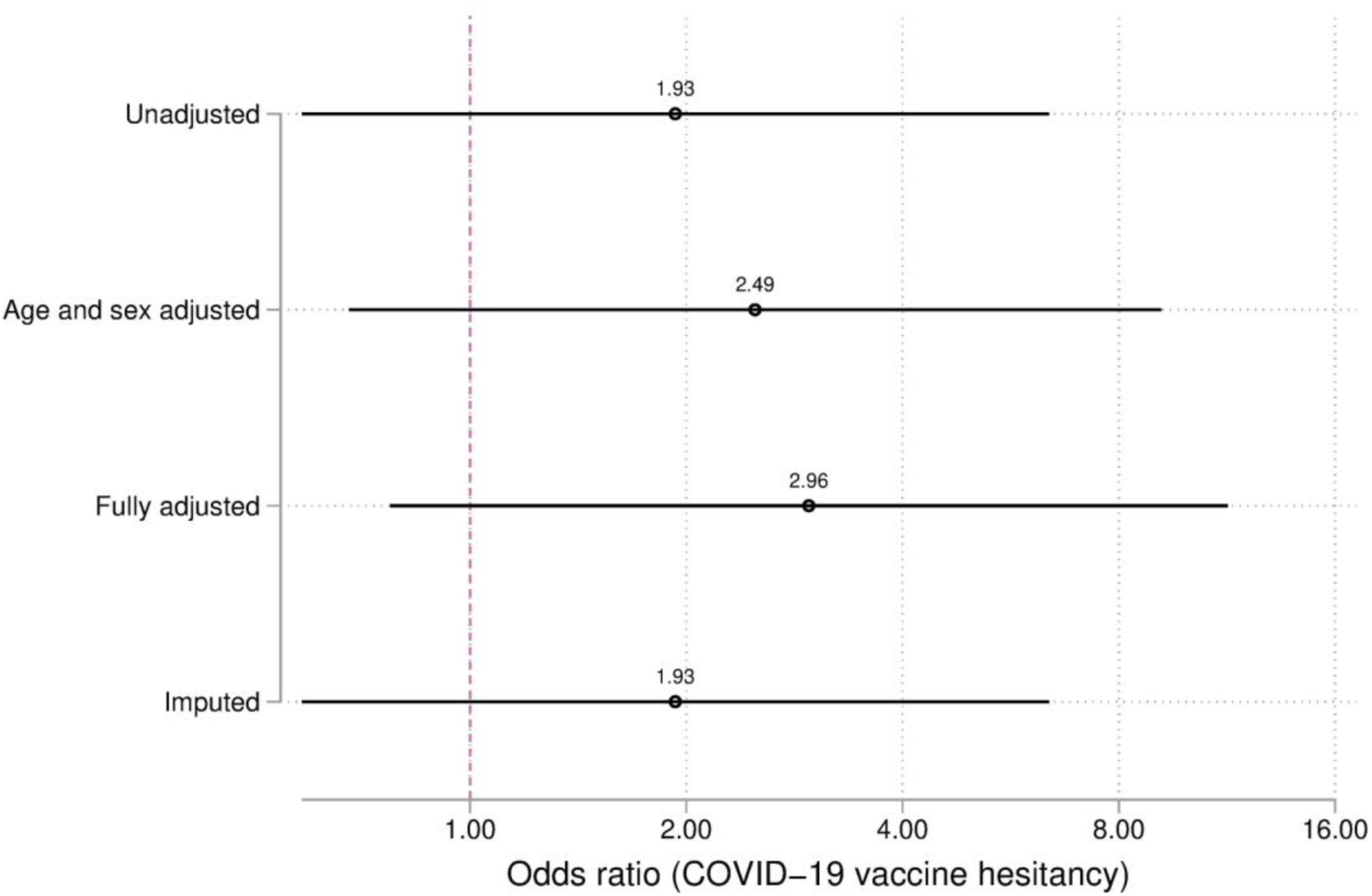
Association between immunosuppressant medication non-adherence and COVID-19 vaccine hesitancy

## Discussion

These global self-reported data characterise both organisational and individual level factors associated with COVID-19 vaccine hesitancy in people with psoriasis. We report COVID-19 vaccine hesitancy in a minority individuals with psoriasis and observe an association between organisational factors (disrupted access to psoriasis care) and hesitancy towards vaccination.

Over one third of participants reported disrupted access to psoriasis care during the pandemic. These individuals were more likely to be younger, of non-white ethnicity, and have shorter duration and more severe psoriasis. A previous single-centre study of 205 psoriasis patients reported lower rates of care disruption (19.5%) but similarly found that patients with severe disease were disproportionately affected^20^. Disruption of care during the pandemic has also been described in other immune-mediated inflammatory diseases. Of 530 participants (the majority of whom had rheumatoid arthritis) from a US-wide questionnaire conducted in March 2020, 20% reported cancelled or postponed appointments^21^. These participants were more likely to have higher disease activity^21^. The COVID-19 Global Rheumatology Alliance (GRA) Patient Experience Survey of 9,300 participants reported later in the pandemic, and found 11% of patients had not been able to communicate with their rheumatologist^22^. There are limited published data in psoriasis or other inflammatory diseases evaluating patient characteristics associated with disrupted care. In our study, participants reporting a negative impact on access to care were also less likely to be taking targeted biologic medications. This may be due to individuals taking biologics requiring clinical interaction with blood test monitoring and disease severity assessments in secondary care, which may have been prioritised during the pandemic. Of note, patients taking standard systemic medications such as methotrexate also require regular blood test monitoring, however, this may take place outside of secondary care settings and these individuals may not require psoriasis specialist review as frequently.

Measuring drug adherence is complex and challenging, with varied rates of adherence reported in psoriasis patients prior to the pandemic. A pre-pandemic systematic review estimated non-adherence rates ranging from 27% to 61% in self-reported studies, with consistently higher adherence to systemic therapies than topical treatments or phototherapy^23^. More recent studies have shown improved adherence amongst those taking systemic therapies^24,25^, one of which found greater adherence to targeted than standard systemic medications^25^. Many intentional and unintentional reasons were reported for non-adherence, including forgetfulness, weak medication-taking routine, concern about side effects or feeling that their psoriasis was under control^24,25^. During the pandemic studies reporting on non**-**adherence to systemic immunosuppressant drugs in rheumatic disease have estimated rates of 15%^26^. We report lower non-adherence rates (17%) than that found in pre-pandemic psoriasis patients but comparable to rheumatic disease patients on systemic medications during the pandemic. The most frequent reported reason for non-adherence in our study was fear of increased infection risk (55%). This was a concern echoed amongst the medical community early in the pandemic prior to the publication of data on COVID-19 mortality rates in individuals taking immunosuppressant therapy^27^. Informed by reassuring data on infection risk, current national and international recommendations support continued immunosuppression during the pandemic, hence communication of these data to patients is vital^28^.

A minority (8%) of our sample reported vaccine hesitancy. Studies indicate that COVID-19 vaccine hesitancy in the global population affects around 26-29% of adults on average, although there is substantial variability between countries^29,30^. Reported hesitancy rates in the UK are around 18%, which are amongst the lowest globally^10^. In our dataset, vaccine hesitancy is generally more prevalent in younger age groups, people of non-white ethnicity and those who live outside the UK. In contrast to other studies we did not find a difference in vaccine hesitancy between genders^10–13,29,30^. Other well described predictors of hesitancy include lower annual income, lower levels of education, higher scores on a COVID-19 conspiracy beliefs scale, worse adherence to COVID-19 guidelines, and lack of influenza vaccine uptake^10–13,29^. Vaccine hesitancy is multi-factorial and context specific, with prior research in the general population (in high income countries) proposing a ‘5C’ model of individual-level drivers encompassing confidence (in vaccine effectiveness/safety), complacency (vaccination felt to not be needed), convenience, risk calculation (risks of infection vs vaccination), and collective responsibility (willingness to protect others by vaccinating one’s self)^9,19^. This provides a framework for exploring individual contributing factors with patients in real-world practice.

There are limited data on vaccine hesitancy among psoriasis patients, particularly since the vaccine programme has begun^16^. A single-centre study of 713 psoriasis patients conducted prior to the COVID-19 vaccine roll-out found that individuals with psoriasis were 32% more willing to be vaccinated than controls who had other skin conditions. In this study, younger age, female sex, higher education level, use of biologic treatment and a history of significant comorbidity correlated with vaccine willingness^16^. The major reasons for vaccine hesitancy were concerns about vaccine safety and efficacy and risk of psoriasis flares^16^. Our findings are in keeping with this, and since our study was conducted after the initiation of the COVID-19 programme, we in part address a limitation of studies focusing on intention alone rather than actual behaviour.

An association between disruption to psoriasis care and COVID-19 vaccine hesitancy is suggested in our dataset (non-statistically significant after adjustment). This is partly mediated by the effects of confounding variables such as age, gender and ethnicity, which associate with both exposure and outcome. Individuals who feel disenfranchised by healthcare services due to disruption to their care may be more likely to be COVID-19 vaccine hesitant. Other studies in the general population indicate that individuals with negative perceptions of doctors have higher vaccine hesitancy, and those with positive healthcare experiences are less likely to be hesitant^12^. Higher expectation for care, as sometimes seen in individuals with more severe disease^31,32^, could also explain the association between disrupted access to care and vaccine hesitancy. Identifying individuals who feel their care has been negatively affected during the pandemic (underserved groups e.g. those of younger age or non-white ethnicity according to our data) will allow us to explore their vaccine concerns (e.g. flares/side effects post-vaccination, vaccine being too new) and ultimately improve vaccine uptake.

The PsoProtect*Me* sample size is large, includes a broad range of ages, psoriasis severity and treatment types. However, the questions are subjective in nature and answered via an online medium which may limit responses to those who are more technologically literate and connected to media. Additionally, psoriasis diagnoses are self-reported and not validated by a clinician. The direction of association cannot be definitively stated due to the cross-sectional nature of this study. The overall impact on care and/or vaccine uptake may have been underestimated since individuals who engage with health surveys may be less likely to feel disenfranchised with healthcare/vaccination services. Social desirability bias may be present, although this is minimised by the anonymous and self-reported nature of this study. We are unable to directly compare our global psoriasis dataset to the general population or other diseases due to a lack of control samples. Our study sample is mostly female (as expected in survey-based studies), from the UK and of white ethnicity, which limits generalisability. Proportionally more patients reported receiving targeted therapies than standard systemic agents, which is not representative of patients receiving systemic therapies and likely indicates ascertainment bias. PsoProtect*Me* was updated one year following its launch to include COVID-19 vaccine hesitancy and access to care questions, hence the current sample may not be representative of the original larger sample^18^.

## Conclusion

This study indicates that a minority of individuals with psoriasis have vaccine hesitancy, which is promising for the uptake of current and future COVID-19 booster vaccines. Vaccine hesitancy was more common in those of younger age and non-white ethnicity. We also identify a substantial disruption to psoriasis care during the pandemic, particularly in those of younger age and non-white ethnicity. Identification of disenfranchised individuals and addressing their concerns regarding the COVID-19 vaccine will help to mitigate risks from the ongoing pandemic.

## Data Availability

Summary level data available on reasonable request.

## Acknowledgements

We are extremely grateful to all the patients who have contributed to PsoProtect*Me*, the professional and patient organizations who supported or promoted PsoProtect*M*e, and Prof Lars Iversen, Prof Nick Reynolds and Prof Joel Gelfand for their valuable input. We would like to acknowledge the following individuals for help with translating the PsoProtect*Me* survey; Dr Haleema Alfailakawi, Dr Wisam Alwan, Dr Rosa Andres Ejarque, Dr Ines Barbosa, Ms Carmen Bugarin Diz, Ms Katarzyna Grys, Dr Mahira Hamdy El Sayed, Mr Tran Hong Truong, Mr Masanori Okuse, Ms Dagmara Samselska, Ms Isabella Tosi, Ms Ya-Hsin Wang. Thank you to the Engine Group UK for their generous creative input and website expertise.

## Notes

**<u>Funding</u>** We acknowledge financial support from the Department of Health via the National Institute for Health Research (NIHR) Biomedical Research Centre based at Guy’s and St Thomas’ NHS Foundation Trust and King’s College London, the NIHR Manchester Biomedical Research Centre and the Psoriasis Association. The views expressed are those of the author(s) and not necessarily those of the NHS, the NIHR, or the Department of Health and Social Care. SKM is funded by a Medical Research Council (MRC) Clinical Academic Research Partnership award (MR/T02383X/1). ND is funded by Health Data Research UK (MR/S003126/1), which is funded by the UK MRC, Engineering and Physical Sciences Research Council; Economic and Social Research Council; Department of Health & Social Care (England); Chief Scientist Office of the Scottish Government Health and Social Care Directorates; Health and Social Care Research and Development Division (Welsh Government); Public Health Agency (Northern Ireland); British Heart Foundation; and Wellcome Trust. ZZNY is funded by a NIHR Academic Clinical Lectureship through the University of Manchester. CEMG is a NIHR Emeritus Senior Investigator and is funded in part by the MRC (MR/101 1808/1). CEMG and RBW are in part supported by the NIHR Manchester Biomedical Research Centre. SML is supported by a Wellcome senior research fellowship in clinical science (205039/Z/16/Z); this research was funded in whole or in part by the Wellcome Trust [205039/Z/16/Z]. For the purpose of Open Access, the author has applied a CC BY public copyright licence to any Author Accepted Manuscript (AAM) version arising from this submission. SML is also supported by Health Data Research UK (grant no. LOND1), which is funded by the UK MRC, Engineering and Physical Sciences Research Council, Economic and Social Research Council, Department of Health and Social Care (England), Chief Scientist Office of the Scottish Government Health and Social Care Directorates, Health and Social Care Research and Development Division (Welsh Government), Public Health Agency (Northern Ireland), British Heart Foundation and Wellcome Trust.

**<u>Conflict of interest disclosures</u>** Nothing to disclose: Dr Bechman, Ms Cook, Dr Dand, Prof. Langan, Dr. Norton, Dr. Tsakok, Dr. Yiu, Dr De La Cruz, Dr. Contreras, Ms. Vesty, Ms. Vincent, Mr. Bola Coker, Ms. Meynell, Dr. Lambert, Prof. Brown, Prof. Naldi. Prof. Barker reports grants and personal fees from Abbvie, grants and personal fees from Novartis, grants and personal fees from Lilly, grants and personal fees from J&J, from null, during the conduct of the study. Prof. Griffiths reports grants and personal fees from AbbVie, grants from Amgen, grants from BMS, grants and personal fees from Janssen, grants from LEO, grants and personal fees from Novartis, grants from Pfizer, grants from Almirall, grants and personal fees from Lilly, grants and personal fees from UCB Pharma, outside the submitted work. Prof. Jullien reports personal fees and non-financial support from Abbvie, personal fees and non-financial support from Novartis, personal fees and non-financial support from Janssen-Cilag, personal fees and non-financial support from Lilly, personal fees and non-financial support from Leo-Pharma, personal fees and non-financial support from MEDAC, personal fees and non-financial support from Celgene, personal fees from Amgen, outside the submitted work. Dr. Capon reports consultancy fees from AnaptysBio, grants from Boheringer-Ingelheim, outside the submitted work. Prof. Bachelez reports personal fees from Abbvie, personal fees from Janssen, personal fees from LEO Pharma, personal fees from Novartis, personal fees from UCB, personal fees from Almirall, personal fees from Biocad, personal fees from Boehringer-Ingelheim, personal fees from Kyowa Kirin, personal fees from Pfizer, outside the submitted work. Prof. Gisondi reports personal fees from Abbvie, Amgen, Eli Lilly, Janssen, Novartis, Pierre Fabre, Sandoz, UCB, outside the submitted work. Dr. Galloway reports personal fees from Abbvie, personal fees from Sanofi, personal fees from Novartis, personal fees from Pfizer, grants from Eli Lilly, personal fees from Janssen, personal fees from UCB, outside the submitted work. Prof. Weinmann has presented talks for Abbvie, Abbott, Bayer, Chiesi, Boehringer Ingelheim, Roche and Merck. Dr. Mason reports personal fees from LEO Pharma and Novartis, outside the submitted work. Ms. Moorhead reports personal fees from Abbvie, personal fees from Celgene, personal fees from Janssen, personal fees from LEO Pharma, personal fees from Novartis, personal fees from UCB, outside the submitted work. Dr. Puig reports grants and personal fees from AbbVie, grants and personal fees from Almirall, grants and personal fees from Amgen, grants and personal fees from Boehringer Ingelheim, personal fees from Bristol Myers Squibb, personal fees from Fresenius-Kabi, grants and personal fees from Janssen, grants and personal fees from Lilly, personal fees from Mylan, grants and personal fees from Novartis, personal fees from Pfizer, personal fees from Sandoz, personal fees from Sanofi, personal fees from Samsung-Bioepis, grants and personal fees from UCB, outside the submitted work. Dr. Mahil reports departmental income from Abbvie, Almirall, Eli Lilly, Janssen-Cilag, Novartis, Sanofi, UCB, outside the submitted work. Dr. Di Meglio reports grants and personal fees from UCB, personal fees from Novartis, personal fees from Janssen, outside the submitted work. Prof. Warren reports grants and personal fees from Abbvie, grants and personal fees from Celgene, grants and personal fees from Eli Lilly, grants and personal fees from Novartis, personal fees from Sanofi, grants and personal fees from UCB|, grants and personal fees from Almirall, grants and personal fees from Amgen, grants and personal fees from Janssen, grants and personal fees from Leo, grants and personal fees from Pfizer, personal fees from Arena, personal fees from Avillion, personal fees from Bristol Myers Squibb, personal fees from Boehringer Ingelheim, outside the submitted work. Prof. Smith reports grants from Abbvie, Sanofi, Novartis, and Pfizer and through consortia with multiple academic partners (psort.org.uk, BIOMAP-IMI.eu), outside the submitted work. Dr. Torres reports grants and personal fees from AbbVie, Almirall, Amgen, Arena Pharmaceuticals, Biogen, Biocad, Boehringer Ingelheim, Bristol-Myers Squibb, Celgene, Eli Lilly, Janssen, LEO Pharma, MSD, Novartis, Pfizer, Samsung-Bioepis, Sandoz, during the conduct of the study. Dr. Waweru is on the Board of the International Federation of Psoriasis Associations who have received grants from Abbvie, Almirall, Amgen, Bristol Meyers Squibb, Boehringer Ingelheim, Celgene, Janssen, Leo Pharma, Eli Lilly, Novartis, Sun Pharma, Pfizer, and UCB, outside the submitted work. Mr. Urmston reports grants from Almirall, grants from Abbvie, grants from Amgen, grants from Celgene, grants from Dermal Laboratories, grants from Eli Lilly, grants from Janssen, grants from LEO Pharma, grants from T and R Derma, grants from UCB, outside the submitted work. Ms. McAteer reports grants from Abbvie, grants from Almirall, grants from Amgen, grants from Celgene, grants from Dermal Laboratories, grants from Eli Lilly, grants from Janssen, grants from LEO Pharma, grants from UCB, grants from T and R Derma, outside the submitted work. Prof. Spuls has done consultancies in the past for Sanofi 111017 and AbbVie 041217 (unpaid), received a departmental independent research grant for TREAT NL registry LeoPharma December 2019; is involved in performing clinical trials with many pharmaceutical industries that manufacture drugs used for the treatment of diseases such as psoriasis and atopic dermatitis, for which financial compensation is paid to the department/hospital; and is chief investigator of the systemic and phototherapy atopic eczema registry (TREAT NL) for adults and children, as well as one of the main investigators of the SECURE-AD registry.

### Competing Interest Statement

Nothing to disclose: Dr Bechman, Ms Cook, Dr Dand, Prof. Langan, Dr. Norton, Dr. Tsakok, Dr. Yiu, Dr De La Cruz, Dr. Contreras, Ms. Vesty, Ms. Vincent, Mr. Bola Coker, Ms. Meynell, Dr. Lambert, Prof. Brown, Prof. Naldi.
Prof. Barker reports grants and personal fees from Abbvie, grants and personal fees from Novartis, grants and personal fees from Lilly, grants and personal fees from J&J, from null, during the conduct of the study.
Prof. Griffiths reports grants and personal fees from AbbVie, grants from Amgen, grants from BMS, grants and personal fees from Janssen, grants from LEO, grants and personal fees from Novartis, grants from Pfizer, grants from Almirall, grants and personal fees from Lilly, grants and personal fees from UCB Pharma, outside the submitted work.
Prof. Jullien reports personal fees and non-financial support from Abbvie, personal fees and non-financial support from Novartis, personal fees and non-financial support from Janssen-Cilag, personal fees and non-financial support from Lilly, personal fees and non-financial support from Leo-Pharma, personal fees and non-financial support from MEDAC, personal fees and non-financial support from Celgene, personal fees from Amgen, outside the submitted work.
Dr. Capon reports consultancy fees from AnaptysBio, grants from Boheringer-Ingelheim, outside the submitted work.
Prof. Bachelez reports personal fees from Abbvie, personal fees from Janssen, personal fees from LEO Pharma, personal fees from Novartis, personal fees from UCB, personal fees from Almirall, personal fees from Biocad, personal fees from Boehringer-Ingelheim, personal fees from Kyowa Kirin, personal fees from Pfizer, outside the submitted work.
Prof. Gisondi reports personal fees from Abbvie, Amgen, Eli Lilly, Janssen, Novartis, Pierre Fabre, Sandoz, UCB, outside the submitted work.
Dr. Galloway reports personal fees from Abbvie, personal fees from Sanofi, personal fees from Novartis, personal fees from Pfizer, grants from Eli Lilly, personal fees from Janssen, personal fees from UCB, outside the submitted work.
Prof. Weinmann has presented talks for Abbvie, Abbott, Bayer, Chiesi, Boehringer Ingelheim, Roche and Merck.
Dr. Mason reports personal fees from LEO Pharma and Novartis, outside the submitted work.
Ms. Moorhead reports personal fees from Abbvie, personal fees from Celgene, personal fees from Janssen, personal fees from LEO Pharma, personal fees from Novartis, personal fees from UCB, outside the submitted work.
Dr. Puig reports grants and personal fees from AbbVie, grants and personal fees from Almirall, grants and personal fees from Amgen, grants and personal fees from Boehringer Ingelheim, personal fees from Bristol Myers Squibb, personal fees from Fresenius-Kabi, grants and personal fees from Janssen, grants and personal fees from Lilly, personal fees from Mylan, grants and personal fees from Novartis, personal fees from Pfizer, personal fees from Sandoz, personal fees from Sanofi, personal fees from Samsung-Bioepis, grants and personal fees from UCB, outside the submitted work.
Dr. Mahil reports departmental income from Abbvie, Almirall, Eli Lilly, Janssen-Cilag, Novartis, Sanofi, UCB, outside the submitted work.
Dr. Di Meglio reports grants and personal fees from UCB, personal fees from Novartis, personal fees from Janssen, outside the submitted work.
Prof. Warren reports grants and personal fees from Abbvie, grants and personal fees from Celgene, grants and personal fees from Eli Lilly, grants and personal fees from Novartis, personal fees from Sanofi, grants and personal fees from UCB|, grants and personal fees from Almirall, grants and personal fees from Amgen, grants and personal fees from Janssen, grants and personal fees from Leo, grants and personal fees from Pfizer, personal fees from Arena, personal fees from Avillion, personal fees from Bristol Myers Squibb, personal fees from Boehringer Ingelheim, outside the submitted work.
Prof. Smith reports grants from Abbvie, Sanofi, Novartis, and Pfizer and through consortia with multiple academic partners (psort.org.uk, BIOMAP-IMI.eu), outside the submitted work.
Dr. Torres reports grants and personal fees from AbbVie, Almirall, Amgen, Arena Pharmaceuticals, Biogen, Biocad, Boehringer Ingelheim, Bristol-Myers Squibb, Celgene, Eli Lilly, Janssen, LEO Pharma, MSD, Novartis, Pfizer, Samsung-Bioepis, Sandoz, during the conduct of the study.
Dr. Waweru is on the Board of the International Federation of Psoriasis Associations who have received grants from Abbvie, Almirall, Amgen, Bristol Meyers Squibb, Boehringer Ingelheim, Celgene, Janssen, Leo Pharma, Eli Lilly, Novartis, Sun Pharma, Pfizer, and UCB, outside the submitted work.
Mr. Urmston reports grants from Almirall, grants from Abbvie, grants from Amgen, grants from Celgene, grants from Dermal Laboratories, grants from Eli Lilly, grants from Janssen, grants from LEO Pharma, grants from T and R Derma, grants from UCB, outside the submitted work.
Ms. McAteer reports grants from Abbvie, grants from Almirall, grants from Amgen, grants from Celgene, grants from Dermal Laboratories, grants from Eli Lilly, grants from Janssen, grants from LEO Pharma, grants from UCB, grants from T and R Derma, outside the submitted work.
Prof. Spuls has done consultancies in the past for Sanofi 111017 and AbbVie 041217 (unpaid), received a departmental independent research grant for TREAT NL registry LeoPharma December 2019; is involved in performing clinical trials with many pharmaceutical industries that manufacture drugs used for the treatment of diseases such as psoriasis and atopic dermatitis, for which financial compensation is paid to the department/hospital; and is chief investigator of the systemic and phototherapy atopic eczema registry (TREAT NL) for adults and children, as well as one of the main investigators of the SECURE-AD registry.

### Funding Statement

We acknowledge financial support from the Department of Health via the National Institute for Health Research (NIHR) Biomedical Research Centre based at Guy's and St Thomas' NHS Foundation Trust and King's College London, the NIHR Manchester Biomedical Research Centre and the Psoriasis Association. The views expressed are those of the author(s) and not necessarily those of the NHS, the NIHR, or the Department of Health and Social Care. SKM is funded by a Medical Research Council (MRC) Clinical Academic Research Partnership award (MR/T02383X/1). ND is funded by Health Data Research UK (MR/S003126/1), which is funded by the UK MRC, Engineering and Physical Sciences Research Council; Economic and Social Research Council; Department of Health & Social Care (England); Chief Scientist Office of the Scottish Government Health and Social Care Directorates; Health and Social Care Research and Development Division (Welsh Government); Public Health Agency (Northern Ireland); British Heart Foundation; and Wellcome Trust. ZZNY is funded by a NIHR Academic Clinical Lectureship through the University of Manchester. CEMG is a NIHR Emeritus Senior Investigator and is funded in part by the MRC (MR/101 1808/1). CEMG and RBW are in part supported by the NIHR Manchester Biomedical Research Centre. SML is supported by a Wellcome senior research fellowship in clinical science (205039/Z/16/Z); this research was funded in whole or in part by the Wellcome Trust [205039/Z/16/Z]. For the purpose of Open Access, the author has applied a CC BY public copyright licence to any Author Accepted Manuscript (AAM) version arising from this submission. SML is also supported by Health Data Research UK (grant no. LOND1), which is funded by the UK MRC, Engineering and Physical Sciences Research Council, Economic and Social Research Council, Department of Health and Social Care (England), Chief Scientist Office of the Scottish Government Health and Social Care Directorates, Health and Social Care Research and Development Division (Welsh Government), Public Health Agency (Northern Ireland), British Heart Foundation and Wellcome Trust.

### Author Declarations

Research approved by KCL research ethics committee (REC ref 20/YH/0135).

## References

1. Stevens S, Pritchard A. IMPORTANT AND URGENT – NEXT STEPS ON NHS RESPONSE TO COVID-19 [Internet]. 2020. Available from: https://www.england.nhs.uk/coronavirus/wp-content/uploads/sites/52/2020/03/urgent-next-steps-on-nhs-response-to-covid-19-letter-simon-stevens.pdf

2. Silva CRDV, Lopes RH, Júnior O de GB, Fuentealba-Torres M, Arcêncio RA, da Costa Uchôa SA. Telemedicine in primary healthcare for the quality of care in times of COVID-19: a scoping review protocol. BMJ Open. 2021 Jul;11(7):e046227. doi: 10.1136/bmjopen-2020-046227.

3. Haldane V, Zhang Z, Abbas RF, Dodd W, Lau LL, Kidd MR, et al. National primary care responses to COVID-19: a rapid review of the literature. BMJ Open. 2020 Dec;10(12):e041622. doi: 10.1136/bmjopen-2020-041622.

4. Thomas SJ, Moreira ED, Kitchin N, Absalon J, Gurtman A, Lockhart S, et al. Safety and Efficacy of the BNT162b2 mRNA Covid-19 Vaccine through 6 Months. N Engl J Med. 2021 Nov 4;385(19):1761–73. doi: 10.1056/NEJMoa2110345.

5. El Sahly HM, Baden LR, Essink B, Doblecki-Lewis S, Martin JM, Anderson EJ, et al. Efficacy of the mRNA-1273 SARS-CoV-2 Vaccine at Completion of Blinded Phase. N Engl J Med. 2021 Nov 4;385(19):1774–85. doi: 10.1056/NEJMoa2113017.

6. Voysey M, Clemens SAC, Madhi SA, Weckx LY, Folegatti PM, Aley PK, et al. Safety and efficacy of the ChAdOx1 nCoV-19 vaccine (AZD1222) against SARS-CoV-2: an interim analysis of four randomised controlled trials in Brazil, South Africa, and the UK. The Lancet. 2021 Jan;397(10269):99–111. doi: 10.1016/S0140-6736(20)32661-1.

7. Office for National Statistics. Coronavirus (COVID-19) latest insights: Vaccines [Internet]. 2021 [cited 2021 Nov 10]. Available from: https://www.ons.gov.uk/peoplepopulationandcommunity/healthandsocialcare/conditionsanddiseases/articles/coronaviruscovid19latestinsights/vaccines

8. Mathieu E, Ritchie H, Ortiz-Ospina E, Roser M, Hasell J, Appel C, et al. A global database of COVID-19 vaccinations. Nat Hum Behav. 2021 Jul;5(7):947–53. doi: 10.1038/s41562-021-01122-8.

9. MacDonald NE. Vaccine hesitancy: Definition, scope and determinants. Vaccine. 2015 Aug;33(34):4161–4. doi: 10.1016/j.vaccine.2015.04.036.

10. Robertson E, Reeve KS, Niedzwiedz CL, Moore J, Blake M, Green M, et al. Predictors of COVID-19 vaccine hesitancy in the UK household longitudinal study. Brain Behav Immun. 2021 May;94:41–50. doi: 10.1016/j.bbi.2021.03.008.

11. Paul E, Steptoe A, Fancourt D. Attitudes towards vaccines and intention to vaccinate against COVID-19: Implications for public health communications. Lancet Reg Health - Eur. 2021 Feb;1:100012. doi: 10.1016/j.lanepe.2020.100012.

12. Freeman D, Loe BS, Chadwick A, Vaccari C, Waite F, Rosebrock L, et al. COVID-19 vaccine hesitancy in the UK: the Oxford coronavirus explanations, attitudes, and narratives survey (Oceans) II. Psychol Med. 2020 Dec 11;1–15. doi: 10.1017/S0033291720005188.

13. Woolf K, McManus IC, Martin CA, Nellums LB, Guyatt AL, Melbourne C, et al. Ethnic differences in SARS-CoV-2 vaccine hesitancy in United Kingdom healthcare workers: Results from the UK-REACH prospective nationwide cohort study. Lancet Reg Health - Eur. 2021 Oct;9:100180. doi: 10.1016/j.lanepe.2021.100180.

14. Salisbury D, Ramsay M, Noakes K. Chapter 14a - COVID-19 - SARS-CoV-2. In: Immunisations Against Infectious Disease (Green Book). Department of Health; 2021.

15. World Health Organisation. WHO SAGE roadmap for prioritizing uses of COVID-19 vaccines in the context of limited supply. 2020.

16. Sotiriou E, Bakirtzi K, Papadimitriou I, Paschou E, Vakirlis E, Lallas A, et al. COVID-19 vaccination intention among patients with psoriasis compared with immunosuppressed patients with other skin diseases and factors influencing their decision. Br J Dermatol. 2021 Jul;185(1):209–10. doi: 10.1111/bjd.19882.

17. PsoProtectMe. Psoriasis Patient Registry for Outcomes, Therapy and Epidemiology of Covid-19 infecTion Me [Internet]. Redcap02. Medstats; [cited 2021 Nov 8]. Available from: https://www.redcap02.medstats.org.uk/redcap/surveys/?s=YTDPT3ADNE

18. Mahil SK, Yates M, Langan SM, Yiu ZZN, Tsakok T, Dand N, et al. Risk-mitigating behaviours in people with inflammatory skin and joint disease during the COVID-19 pandemic differ by treatment type: a cross-sectional patient survey*. Br J Dermatol. 2021 Jul;185(1):80–90. doi: 10.1111/bjd.19755.

19. Betsch C, Schmid P, Heinemeier D, Korn L, Holtmann C, Böhm R. Beyond confidence: Development of a measure assessing the 5C psychological antecedents of vaccination. Angelillo IF, editor. PLOS ONE. 2018 Dec 7;13(12):e0208601. doi: 10.1371/journal.pone.0208601.

20. Ninosu N, Roehrich F, Diehl K, Peitsch WK, Schaarschmidt M-L. Psoriasis care during the time of COVID-19: real-world data on changes in treatments and appointments from a German university hospital. Eur J Dermatol. 2021 Apr;31(2):183–91. doi: 10.1684/ejd.2021.4016.

21. Michaud K, Wipfler K, Shaw Y, Simon TA, Cornish A, England BR, et al. Experiences of Patients With Rheumatic Diseases in the United States During Early Days of the COVID-19 Pandemic. ACR Open Rheumatol. 2020 Jun;2(6):335–43. doi: 10.1002/acr2.11148.

22. Hausmann JS, Kennedy K, Simard JF, Liew JW, Sparks JA, Moni TT, et al. Immediate effect of the COVID-19 pandemic on patient health, health-care use, and behaviours: results from an international survey of people with rheumatic diseases. Lancet Rheumatol. 2021 Oct;3(10):e707–14. doi: 10.1016/S2665-9913(21)00175-2.

23. Thorneloe RJ, Bundy C, Griffiths CEM, Ashcroft DM, Cordingley L. Adherence to medication in patients with psoriasis: a systematic literature review. Br J Dermatol. 2013 Jan;168(1):20–31. doi: 10.1111/bjd.12039.

24. Hambly R, Kelly A, Gilhooley E, Nic Dhonncha E, Murad A, Hughes R, et al. Medication adherence among patients with psoriasis on traditional systemic and biologics treatment. Br J Dermatol [Internet]. 2018 Jan;178(1). doi: 10.1111/bjd.15856.

25. Thorneloe RJ, Griffiths CEM, Emsley R, Ashcroft DM, Cordingley L, Barker J, et al. Intentional and Unintentional Medication Non-Adherence in Psoriasis: The Role of Patients’ Medication Beliefs and Habit Strength. J Invest Dermatol. 2018 Apr;138(4):785–94. doi: 10.1016/j.jid.2017.11.015.

26. Rebić N, Park J, Garg R, Ellis U, Kelly A, Davidson E, et al. A rapid review of medication taking (‘adherence’) among patients with rheumatic diseases during the COVID-19 pandemic. Arthritis Care Res. 2021 Jul 5;acr.24744. doi: 10.1002/acr.24744.

27. MacKenna B, Kennedy NA, Mehkar A, Rowan A, Galloway J, Mansfield KE, et al. Risk of severe COVID-19 outcomes associated with immune-mediated inflammatory diseases and immune modifying therapies: a nationwide cohort study in the OpenSAFELY platform [Internet]. Infectious Diseases (except HIV/AIDS); 2021 Sep. doi:10.1101/2021.09.03.21262888.

28. Gelfand JM, Armstrong AW, Bell S, Anesi GL, Blauvelt A, Calabrese C, et al. National Psoriasis Foundation COVID-19 Task Force Guidance for Management of Psoriatic Disease During the Pandemic: Version 1. J Am Acad Dermatol. 2020 Dec;83(6):1704– 16. doi: 10.1016/j.jaad.2020.09.001.

29. Lazarus JV, Ratzan SC, Palayew A, Gostin LO, Larson HJ, Rabin K, et al. A global survey of potential acceptance of a COVID-19 vaccine. Nat Med. 2021 Feb;27(2):225– 8. doi: 10.1038/s41591-020-1124-9.

30. Neumann-Böhme S, Varghese NE, Sabat I, Barros PP, Brouwer W, van Exel J, et al. Once we have it, will we use it? A European survey on willingness to be vaccinated against COVID-19. Eur J Health Econ. 2020 Sep;21(7):977–82. doi: 10.1007/s10198-020-01208-6.

31. Bhutani T, Wong JW, Bebo BF, Armstrong AW. Access to health care in patients with psoriasis and psoriatic arthritis: data from National Psoriasis Foundation survey panels. JAMA Dermatol. 2013 Jun;149(6):717–21. doi: 10.1001/jamadermatol.2013.133.

32. Duvetorp A, østergaard M, Skov L, Seifert O, Tveit KS, Danielsen K, et al. Quality of life and contact with healthcare systems among patients with psoriasis and psoriatic arthritis: results from the NORdic PAtient survey of Psoriasis and Psoriatic arthritis (NORPAPP). Arch Dermatol Res. 2019 Jul;311(5):351–60. doi: 10.1007/s00403-019-01906-z.

