## Supplementary File for "Vaccine hesitancy and access to psoriasis care in the COVID-19 pandemic: findings from a global patient-reported cross-sectional survey"

**Supporting Information**

**Appendix S1**

**Imputation methodology**

Due to missing data (Supplementary Table S1), age, gender, ethnicity, and treatment variables were imputed using complete data from the individuals in the population who had completed questionnaires (n=4361). A multiple imputation model was employed, using self-reported complete data from the 3,338 individuals in the population who had completed the questionnaire but had not answered sections on impact of COVID-19 on care and vaccine uptake, and complete data from the 655 individuals who had answered sections on impact of COVID-19 on care and vaccine uptake. There were differences in baseline demographics between these two cohorts (Supplementary Table S2). Predictive mean matching was performed to impute the missing variables. The data were imputed using multivariate sequential imputation, using chained equations. Firstly, all missing values were filled in by simple random sampling with replacement from the observed values. The imputation was 20 cycles, where at the end of the cycle one imputed dataset was created and the process was repeated to create 20 imputed datasets. The 20 datasets were combined using Rubin’s rules, therefore the estimates and standard errors presented are the combined ones.

**Table S1.** Missing data

|  | **Missing (n)** | **Present (n)** | **Total (n)** |
| --- | --- | --- | --- |
| **Age** | 144 (18.0%) | 658 | 802 |
| **Gender** | 142 (17.7%) | 660 | 802 |
| **Ethnicity** | 147 (18.3%) | 655 | 802 |
| **Smoking Status** | 166 (20.7%) | 636 | 802 |
| **Treatment** | 58 (7.2%) | 744 | 802 |
| **BMI** | 200 (24.9%) | 602 | 802 |
| **Vaccine Hesitancy** | 47 (5.9%) | 755 | 802 |

**Table S2.** Demographic and disease specific characteristics of participants included in main analysis (answered questions on access to care and vaccine hesitancy) and in the remaining cohort.

|  | **Participants included in the analysis** | **Remaining cohort, not in analysis.** | **p-value** |
| --- | --- | --- | --- |
|  | N=655/802 | N=3,338/3,559 |  |
| **Age** | 51.0 (37.0-61.0) | 46.0 (35.0-57.0) | <0.001 |
| **Gender** |  |  | 0.56 |
| Female | 430 (65.6%) | 2,274 (66.8%) |  |
| Male | 225 (34.4%) | 1,129 (33.2%) |  |
| **BMI** | 26.6 (23.1-30.7) | 26.8 (23.4-31.2) | 0.18 |
| **Smoking Status** |  |  | 0.79 |
| Current tobacco smoker | 81 (12.8%) | 408 (13.7%) |  |
| Former tobacco smoker | 222 (35.2%) | 1,060 (35.6%) |  |
| Never smoked | 328 (52.0%) | 1,511 (50.7%) |  |
| **Ethnicity** |  |  | 0.047 |
| White | 579 (88.4%) | 2,802 (83.9%) |  |
| Black | 5 (0.8%) | 60 (1.8%) |  |
| Asian | 43 (6.6%) | 284 (8.5%) |  |
| Hispanic/Latinx | 12 (1.8%) | 76 (2.3%) |  |
| Other | 16 (2.4%) | 116 (3.5%) |  |
| **Country** |  |  | 0.009 |
| United Kingdom | 481 (73.9%) | 2,300 (68.3%) |  |
| North America | 61 (9.4%) | 378 (11.2%) |  |
| Other Europe | 65 (10.0%) | 339 (10.1%) |  |
| Rest of World | 44 (6.8%) | 351 (10.4%) |  |
| **Duration of Psoriasis (Years)** | 28.0 (14.0-42.0) | 23.0 (11.0-35.0) | <0.001 |
| **Psoriasis Severity** |  |  | 0.074 |
| Mild | 406 (62.5%) | 1,936 (60.8%) |  |
| Moderate | 162 (24.9%) | 730 (22.9%) |  |
| Moderate-Severe | 55 (8.5%) | 380 (11.9%) |  |
| Severe | 27 (4.2%) | 138 (4.3%) |  |
| **Treatment** |  |  | 0.49 |
| No Systemic Therapy | 360 (58.4%) | 1,677 (56.1%) |  |
| Standard Systemic Therapy | 89 (14.4%) | 479 (16.0%) |  |
| Targeted Therapy | 167 (27.1%) | 834 (27.9%) |  |
